# Genetic risk for Alzheimer’s disease, cognition and Mild Behavioral Impairment in healthy older adults

**DOI:** 10.1101/2020.05.13.20100800

**Authors:** Byron Creese, Helen Brooker, Dag Aarsland, Anne Corbett, Clive Ballard, Zahinoor Ismail

## Abstract

**BACKGROUND:** Mild Behavioral Impairment (MBI) is a neuropsychiatric syndrome describing later-life emergent apathy, mood/anxiety symptoms, impulse dyscontrol, social inappropriateness and psychosis that are not attributable to psychiatric diagnoses. MBI is an at-risk state for incident cognitive decline and dementia, and is associated with dementia biomarkers including Aβ and neurofilament light. Thus, MBI may be an early clinical marker of neurodegenerative disease. In this study, we hypothesized that stratification by MBI in a cognitively normal sample would moderate the signal between Alzheimer’s disease (AD) genetic risk and cognition.

**METHODS:** Genetic, cognitive and MBI data was available for 3,126 PROTECT study participants over 50 without dementia. A general cognitive composite score was constructed based on scores on paired associates learning, digit span, self-ordered search and verbal reasoning. MBI was assessed using the MBI Checklist. Polygenic scores for AD were split by tertile (representing low, medium and high risk) and the sample was stratified by MBI into those with no symptoms and those with any symptoms.

**RESULTS:** AD genetic risk was associated with poorer cognition in the MBI strata only (MBI: F(2,1746)=4.95, p=0.007; no MBI: F(2,1366)=0.72, p=0.49). The mean difference between low and high genetic risk groups was significant (p=0.005) and the standardised effect size in the MBI sample was higher than in the whole sample.

**CONCLUSIONS:** These findings justify MBI screening to enrich samples with at-risk individuals, and underscore the importance of late-life neuropsychiatric symptoms in cognitive ageing.

## Background

Worldwide, the number of people with dementia is expected to rise to 150 million by 2050. Recent years have been marked by a number of high profile failures of disease modifying therapies and it is now widely recognised that identification of people in the very earliest stages of dementia is a key priority for clinical trials of new treatments, and ultimately for clinical practice [1]. Genetic predictors of cognitive decline and dementia have been the subject of considerable focus in recent years. These predictors include not only apolipoprotein E (APOE) status but also polygenic risk scores (PRS). PRS are the sum of AD risk alleles carried by an individual weighted by effect size, and therefore capture more genetic risk than APOE alone. The ultimate goal of this work is the identification of low cost early marker of neurodegenerative disease. As an important first step, numerous studies have shown that AD genomic markers predict AD and mild cognitive impairment (MCI) case/control status, as well as progression to AD among MCI cases [2–7]. However, the association between AD genetic risk and objectively measured cognition in non-dementia samples is less consistent. Although at least nine studies have examined this question, four have reported a no link [8–15]. There are a number of methodological differences which likely explain these discrepancies, including the sensitivity of cognitive outcome measures and the number of risk alleles included in PRS calculation. One additional challenge is the complex etiology of cognition in older adults, which is not solely accounted for by genetic risk for neurodegeneration or neuropsychological profile. Because of the ease and low cost of genetic analysis, and promising initial findings, it is logical to explore strategies that may enhance sensitivity.

One study found that stratification based on Aβ positive positron emission tomography (PET) scans unmasked an association between AD PRS and poorer memory and executive function [12]. That neuropathological markers of AD moderate the association between AD genetic risk and neuropsychological measures is not surprising, but does offer proof of principle of sample enrichment, informing our study. In order to add value to a genetic screen, such sample enrichment should be low cost and scalable to large populations. Scalability is not achievable with PET imaging due to cost and barriers to access, but there is evidence that later-life emergent neuropsychiatric symptoms (NPS), described by the validated syndrome, Mild Behavioral Impairment (MBI), may represent such a screening tool. MBI is a neurobehavioral syndrome proposed by an Alzheimer’s Association consensus group to describe a risk state for cognitive decline and dementia in order to facilitate earlier dementia detection [16]. The MBI syndrome covers late-life emergent apathy, mood/anxiety symptoms, impulse dyscontrol, social inappropriateness, and psychotic symptoms, and is common and easily measured in the general population [17]. Moreover, MBI is associated with progressive cognitive decline in individuals without significant cognitive impairment and a shorter time to dementia in individuals with normal cognition or MCI at baseline [18–22]. Crucially, MBI has recently been shown to associate with known dementia biomarkers. In a sample of cognitively normal older adults, MBI score was associated with a greater burden of PET amyloid, suggesting it is a novel neuropsychiatric marker of preclinical disease [23]. In a sample of non-demented older adults with normal cognition and MCI, MBI was associated with faster accumulation of neurofilament light, which is a marker of axonal loss, and there is preliminary evidence that MBI is associated with AD PRS [24,25]. On the basis of this evidence linking MBI to the neurobiology of dementia, MBI is an attractive candidate tool to enrich samples with individuals at greater risk of dementia. To examine this application of MBI, we tested whether the relationship between AD genetic risk and cognition was moderated by MBI symptoms in a cohort of older adults without dementia. We hypothesised AD genetic risk would be associated with poorer cognition and that this relationship would be strongest among people with MBI symptoms.

## Method

### Participants

Data from 4,591 participants taking part in the Platform for Research Online to Investigate Genetics and Cognition in Aging (PROTECT) study were analysed in this study (REC reference 13/LO/1578). Written informed consent was obtained from all participants and proxy informants. PROTECT is a UK-based online participant registry which tracks the cognitive health of older adults. Inclusion criteria for enrolling in PROTECT are 1) >=50 years old; 2) no diagnosis of dementia; and 3) access to a computer and internet. This sample is a subset of PROETCT study participants who also provided a saliva sample for genotyping, completed cognitive testing and had a proxy informant available to complete the MBI Checklist (MBI-C, further detail is presented below).

### Assessment of Cognition

Cognitive performance was assessed via a battery of four tests (Table 1). Individual performance across cognitive tests is known to be correlated, and for this study we analysed a general cognitive composite based on factor analysis of the battery. This latent construct, capturing general cognitive ability, is a well-documented feature of cognition [26]. To capture general cognitive ability in this sample, a composite score was calculated by computing the first unrotated principal component of the cognitive battery. The variance in total cognitive test score explained by the first principal component was 48.4% and the factor loadings were 0.53 (paired associated learning), 0.5 (digit span), 0.46 (self-ordered search) and 0.51 (verbal reasoning). This finding is comparable to recent reports in an analysis of over 300,000 individuals across multiple cohorts [27]. In the present study, lower cognitive composite score was associated higher Informant Questionnaire on Cognitive Decline in the Elderly (IQCODE) score, which indicates a negative change in cognition over 10 years via a questionnaire observed by a proxy informant (β[SE]=-0.16[0.04], p=2.72×10^−5^), and greater impairment in instrumental activities of daily living (IADL) as measured by the Minimum Data Set-Home Care IADL scale (β[SE]=-0.23 [0.04], p=1.09×10^−9^), providing validation that the cognitive construct is meaningful in the context of our sample [28,29].

**Table 1.**
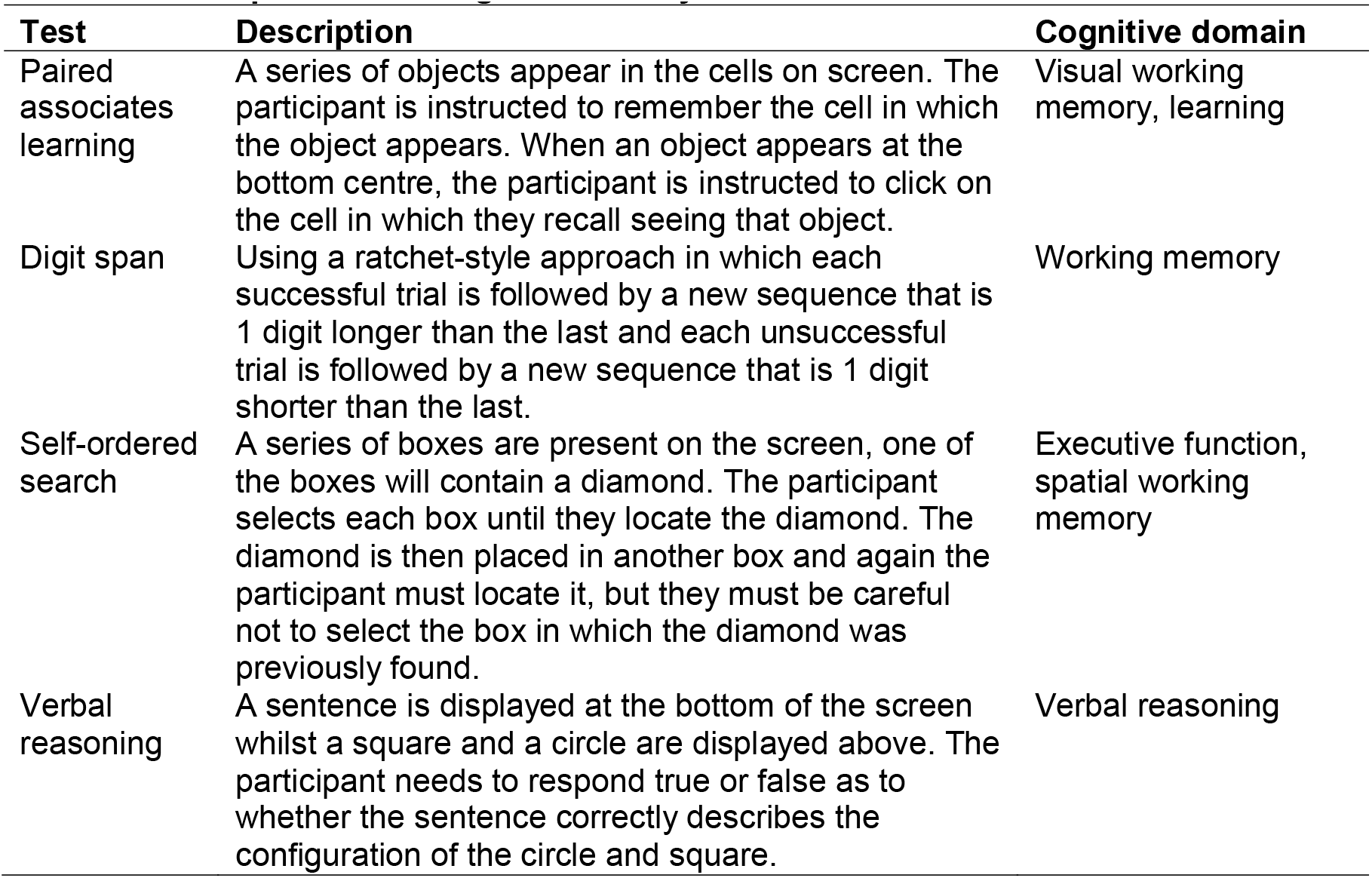
Description of the cognitive battery used

### Assessment of neuropsychiatric symptoms

Neuropsychiatric symptoms were operationalized in the MBI framework using the MBI Checklist (MBI-C). The MBI-C is a validated tool designed specifically for capturing MBI symptoms and, in this study, was completed by a proxy informant who knew the participant well for at least 10 years [17,30–32]. The scale consists of 34 questions covering the full range of MBI domains (apathy, mood/anxiety symptoms, impulse dyscontrol, social inappropriateness, and psychotic symptoms). Each question is rated on a scale of 0 (not present) to 3 (severe). The MBI-C mandates that a symptom must be present for at least 6 months and represent a change from longstanding behaviour in order to be rated as present. This approach facilitates differentiation of MBI symptoms from transient neuropsychiatric symptoms and reactive conditions due to medical and environmental precipitants, in order to better reflect the new onset symptomatology seen in neurodegenerative disease [33]. In this study participants were classified as having any symptoms of MBI (MBI-C total score >0) or having no MBI symptoms (MBI-C total score=0) due to the strong positive skew of the MBI-C data (i.e. ∼50% of respondents can be expected to score zero) [17]. Any participants with the following medical conditions were excluded (all derived from self-report responses to the question “Have you ever been diagnosed with one or more of the following even if you don’t have it currently?”: mild cognitive impairment (n=70), stroke (n=375), Parkinson’s disease (n=47), depression (n=592), mania/bipolar/manic depression (n=152), anxiety/generalized anxiety disorder (n=12), panic attacks (n=3), anorexia nervosa (n=135), bulimia nervosa (n=14), psychological overeating/binge eating (n=1), autism/Asperger’s/autistic spectrum disorder (n=1), attention deficit disorder (n=1). The questionnaire also covered schizophrenia/other psychotic illnesses, personality disorders and social anxiety/phobia but no participants reported having these. We also excluded anyone who scored>=13 on the Patient Health Questionnaire 9 (PHQ9), indicating a probably current major depressive episode (n=253). These exclusions were applied in order to reflect the Alzheimer’s Association MBI diagnostic criteria stipulation that symptoms cannot be better explained by a pre-existing medical or psychiatric condition [16]; 1,463 individual participants met at least one of these exclusion criteria.

The binary coding of the MBI-C is supported by recent data showing that in cognitively normal people, a score >0 on the MBI-C (i.e. the presence of any symptoms of any severity)was associated with worse cognitive performance (both at baseline and over one year) on a range of tests, with a score of >8 being associated with the worst performance [19].

### Genetic data QC and AD polygenic risk score calculation

Using PLINK, SNP and individual quality control exclusions were applied to genotype data (minor allele frequency (MAF) ≤1%, Hardy Weinberg equilibrium p<10^−5^, SNP and individual missingness >2%, mean heterozygosity ±3SD, chip-gender mismatches, non-European ancestry (derived from genetic principal components calculated in PLINK and projected onto HapMap phase 3 populations, see Supplementary Material), related/duplicate (pi-hat >0.2) samples). Phasing (EAGLE2) and imputation (PBWT) was done via the Sanger Imputation Service using the Haplotype Reference Consortium (r1.1) reference panel. SNPs with imputation quality (INFO) score ≥0.8 and MAF ≥0.01 were retained, leaving 6,782,377 available for analysis. IGAP AD GWAS was used to calculate PRS using PRSice (clumped using 250kb windows and r^2^>0.1) [34,35]. Previous data suggests a wide range of inclusion thresholds for PRS calculation, from only GWAS significant SNPs to many thousands of SNPs [2,3,8–15]. In this study, we opted for two AD GWAS SNP inclusion thresholds (PT). The first, 1×10^−5^(93 SNPS), was chosen because it was the most strongly associated with a family history of dementia (β=0.07, SE=0.01, p=4.54×10^−13^) in the PROTECT study and the second was all IGAP SNPs (i.e.PT=1). AD PRS at PT=1 was also associated with family history status (β=0.02, SE=0.01, p=0.01). PRS without the APOE locus (chr19:44,00,000-46,500,000) were also calculated.

### Statistical analysis

PRS were standardized before analysis. PRS were split by tertile with the bottom 1/3 representing low AD genetic risk, the middle 1/3 medium genetic risk and the top 1/3 high genetic risk. The cognitive composite score was normalised for age, sex and education using a linear regression models and the standardised residuals used in subsequent analysis. The association between AD genetic risk and cognition was first tested by ANCOVA in the whole sample at both *P_T_*. The sample was then stratified by MBI status and the dependent variable of cognitive composite was analysed by ANCOVA with AD genetic risk group (three level factor) and the first four ancestry principal components as covariates. The effect size for the overall ANCOVA was expressed by partial eta-squared (η^2^). Tukey post-hoc test was used to undertake planned pairwise comparisons between levels of AD genetic risk, and effect sizes for mean differences between these groups were expressed by Cohen’s d. ANCOVA assumptions were checked by examining residual plots and checking for outliers. Given that the two AD PRS are not independent, and a Bonferroni correction would therefore be overly conservative, a family-wise error rate of 0.025 was applied (to reflect tests on the two MBI symptom groups). Post-hoc comparisons between the three AD genetic risk groups were Bonferroni adjusted. All statistical analysis was performed in R.

## Results

### Participant characteristics

Following medical and psychiatric history exclusions, 3,126 participants were available for analysis. Participant characteristics by cognitive group are shown in Table 2. There was no difference in age, sex or education level between the MBI and no MBI strata (age: t=0.41, df=3034.4, p=0.68; sex: X=0.035, df=1, p=0.85; education level: X=6.65, df=5, p=0.25) but,as expected, the MBI group did have a lower cognitive score (t=2.87, df=2948.1, p=0.004,Cohen’s *d*=0.1).

**Table 2.** Participant characteristics by cognitive group

|  | MBI strata |  |  |  | P |
| --- | --- | --- | --- | --- | --- |
|  | No Symptoms (MBI-C=0) |  | MBI Symptoms (MBI-C>0) |  |  |
| N (%) | 1373 | 44 | 1753 | 56 |  |
| Age (mean, sd) | 63.6 | 6.5 | 63.5 | 7.03 | 0.68* |
| Sex (n, %) |  |  |  |  |  |
| Male | 314 | 23 | 407 | 23 | 0.85† |
| Female | 1059 | 77 | 1346 | 77 |  |
| Education level (n, %) |  |  |  |  |  |
| Secondary education | 149 | 11 | 227 | 13 | 0.65† |
| Post-secondary education | 146 | 11 | 208 | 12 |  |
| Vocational qualification | 270 | 20 | 326 | 19 |  |
| Undergraduate degree | 508 | 37 | 605 | 35 |  |
| Post graduate degree | 235 | 17 | 315 | 18 |  |
| Doctoral degree | 65 | 5 | 72 | 4 |  |
| Cognitive composite (mean, sd)‡ | 0.26 | 1.2 | 0.14 | 1.18 | 0.004* |

Abbreviations: MBI, Mild Behavioral Impairment; MBI-C, Mild Behavioral Impairment Checklist; sd, Standard Deviation
\*p-value of t-test
†p-value of chi-square test
‡Cognitive composite is the first unrotated principal component derived from scores on paired associates learning, digit span, self-ordered search and verbal reasoning

### Relationship betweenAD PRS,cognition and neuropsychiatric symptoms

In the whole sample analysis, AD genetic risk at *P_T_*=1 was associated with a lower mean cognitive composite score (F(2,3119)=3.93, p=0.02, partial η^2^=0.003) but not at the more conservative PRS at *P_T_*=1×10^−5^ (F(2,3119)=1.11, p=0.33). Pairwise comparisons showed that the mean difference in cognitive score for the high genetic risk and low genetic risk groups was statistically significant but there were no differences between the other groups(mean difference: −0.15, p=0.02, Cohen’s d=0.13, Table 3). Linear regression using the untransformed AD PRS (i.e. as a normally distributed continuous level variable rather than tertiles) produced a similar result (β[SE]=-0.05[0.02], p=0.03, with analysis at *P_T_*=1×10^−5^ remaining non-significant). Accordingly, the rest of the analysis was only conducted on AD PRS at *P_T_*=1.

In stratified analysis, shown in Table 3and Figure 1, the association between AD genetic risk and cognition persisted but only in those with MBI symptoms, where the effect size was larger than in the whole sample analysis (F(2,1746)=4.95, p=0.007, partial η^2^=0.006). In those with no MBI symptoms there was no association between AD genetic risk and cognition (F(2,1366)=0.72, p=0.49). Pairwise comparisons in the MBI symptom sample showed that mean difference in cognitive scores between the high and low genetic risk groups was again statistically significant, with a larger effect size than in the whole sample analysis, Cohen’s d increased from 0.13 to 0.19 (mean difference: −0.22, p=0.005). The association remained statistically significant after removing the APOE locus, although the effect size was attenuated (F(2,1746)=3.23, p=0.04, partial η^2^=0.002; mean difference between high and low genetic risk groups: −0.17, p=0.03, Cohen’s *d*=0.14).

**Figure 1:**
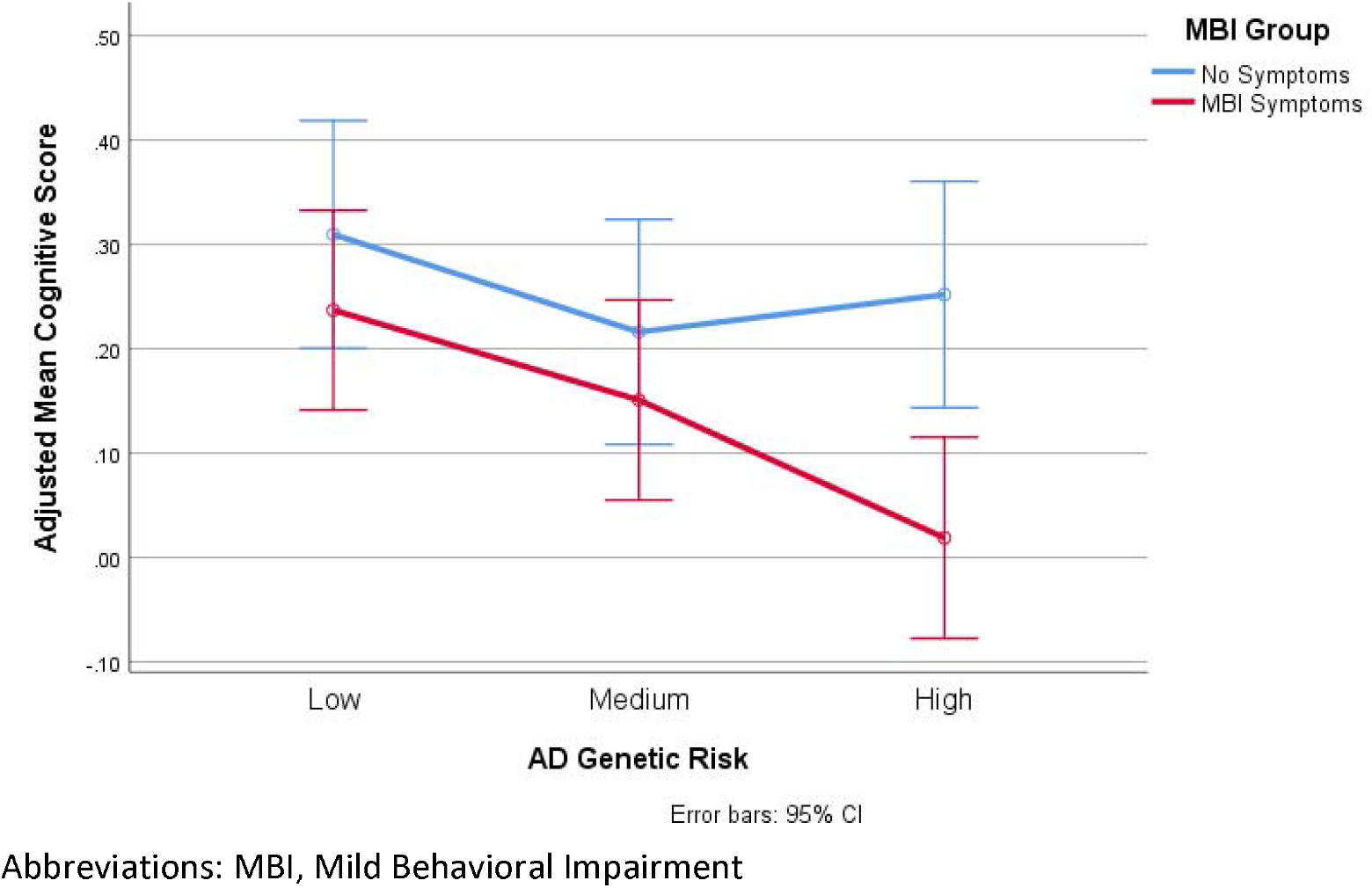
Plot of adjusted mean cognitive composite score by AD genetic risk group(PT=1) stratified by MBI symptom grouping.

**Table 3.** Mean differences in cognitive scores by AD genetic risk group stratified by MBI symptoms

|  | ANCOVA |  |  |  | Cognitive test score by AD genetic risk |  |  |  | Pairwise comparisons* |  |  |
| --- | --- | --- | --- | --- | --- | --- | --- | --- | --- | --- | --- |
|  | N | F | df | P | Group | n | Adjusted Mean | SD |  | P | Cohen's d |
| Whole sample | 3216 | 3.93 | 2,3119 | 0.02 | High | 1037 | 0.12 | 1.15 | High vs Low | 0.02 | 0.13 |
|  |  |  |  |  | Medium | 1049 | 0.18 | 1.21 | High vs Middle | 0.21 | 0.05 |
|  |  |  |  |  | Low | 1040 | 0.27 | 1.19 | Low vs Middle | 0.51 | 0.07 |
| MBI Symptoms | 1753 | 4.95 | 2,1746 | 0.007 | High | 579 | 0.02 | 1.07 | High vs Low | 0.005 | 0.19 |
|  |  |  |  |  | Medium | 586 | 0.15 | 1.25 | High vs Middle | 0.45 | 0.11 |
|  |  |  |  |  | Low | 588 | 0.24 | 1.21 | Low vs Middle | 0.13 | 0.07 |
| No symptoms | 1373 | 0.72 | 2,1366 | 0.49 | High | 458 | 0.25 | 1.23 | - | - | - |
|  |  |  |  |  | Medium | 463 | 0.22 | 1.16 | - | - | - |
|  |  |  |  |  | Low | 452 | 0.31 | 1.17 | - | - | - |

Abbreviations: MBI, Mild Behavioral Impairment.
\*Post-hoc pairwise comparison computed by Tukey test (p-values are Bonferroni adjusted).
∴ pairwise comparisons not undertaken as ANCOVA was not significant.

### Sensitivity analysis

In our primary analysis we adopted a binary coding for MBI symptoms (none vs. any) because of evidence that even individuals with low level symptoms have an increased risk of cognitive decline. There are no accepted cut points on the MBI-C in cognitively normal community samples so, post-hoc, we examined the association between AD PRS, cognitive impairment and MBI using an MBI-C cut point of ≥3. For context, this means that the MBI group now contained any individual who had experienced at least one ‘severely’ rated symptom or multiple less severe symptoms, while the no MBI group contained all other participants; this resulted in 2,020 in the no symptoms group and 1,106 in the MBI group. The associations observed in the main analysis were sustained with a similar effect size (MBI symptoms: F(1,1099)=3.01, p=0.048, partial η^2^=0.005; No symptoms:F(1,2013)=1.79,p=0.17) and the magnitude of the differences between the high and low genetic risk groups in the MBI symptom sample was the same(mean difference: −0.20, p=0.047, Cohen’s d=0.19).

## Discussion

To our knowledge, these findings are the first demonstration that sample stratification on neuropsychiatric symptoms (here assessed in the MBI framework) enhances the effect size of the association between AD genetic risk and cognition in a sample of cognitively normal older adults. The association was present in our study using a PRS based on all available SNPs from AD GWAS (*P_T_*=1) and remained after exclusion of the APOE locus, albeit at a diminished effect size. Taken together we conclude that APOE is driving much of the signal observed in this study, consistent with previous reports, but that non-APOE SNPs also play a role in late life cognition. More broadly, our findings from this sample, which is independent from previous reports, support a role for AD genetic risk, beyond APOE, in cognition among older adults without dementia, an important finding as there is not unanimity in previous literature [8–11,13–15].

A recent well phenotyped large study of older adults found that AD PRS which included all SNPs (PT=1) was not associated with cognition in adults but APOE was [14]. Genetic risk for AD, including APOE and other SNPs, has been shown to be pleiotropic but it is notable that a previous study found the effect of APOE on cognition to be stronger in older adults relative to earlier in life [36,37]. Older samples will likely contain a larger number of individuals in prodromal or preclinical disease so a stronger effect could be reasonably expected for this reason. Similarly, we propose that stratifying on MBI, even among cognitively normal older adults, has the same effect and defines a cognitive phenotype which is ‘closer’ to Alzheimer’s disease. This in turn would create better power to detect associations with those AD SNPs with smaller effect sizes, thus explaining the discrepancy between our finding and this other recent work.

The notion that neuropsychiatric symptoms (i.e. MBI) may be an early marker of dementia is somewhat counterintuitive for a group of diseases that are primarily conceptualised as cognitive disorders. This cognocentric approach does not necessarily reflect the history of AD. Auguste D., the index patient described by Alois Alzheimer presented to hospital with emotional dysregulation and suspiciousness, followed by cognitive decline [33,38–40]. Several longitudinal studies support MBI emerging in advance of cognitive symptoms or increasing risk of incident cognitive decline and dementia [18,20,21,41]. The distinction between MBI and other neuropsychiatric symptoms as risk factors vs. early markers has been a topic of recent debate [42,43]. However, new data in cognitively normal people showing that MBI symptoms are associated with higher amyloid beta burden [23] and faster accumulation of neurofilament light [24] support the view of MBI as an early clinical marker. The implication is therefore that stratification on MBI symptoms enriches samples for individuals with preclinical or prodromal disease, creating a more etiologically homogenous sample. Deeper phenotyping including fluid imaging and longitudinal follow up with detailed neuropsychology and clinical outcomes will be required to confirm this hypothesis, which could have important implications for clinical trials where cohort heterogeneity has been identified as a major concern [44].

The operationalization of MBI is worth some discussion. A key strength of this study is the exclusion of pre-existing psychiatric conditions and use of the specific MBI-C tool, both of which provide more confidence that our findings are due to later-life emergent mild NPS rather than longstanding clinically significant psychiatric diagnoses. However, establishing appropriate cut points on the MBI-C is a matter of ongoing research. Our findings, including our post-hoc analysis, suggest that relatively mild symptoms are important to consider and that the likely optimal cut point on the MBI-C would lie somewhere between 2 and 8. This is supported by previous research in showing that low level symptoms of MBI as well as the more severe symptoms are associated with cognitive decline in cognitively healthy individuals [19]. Relating to this, a limitation to our study is the proxy completion of MBI-C via remote questionnaire completion, which could have led to some misclassifications, although raters in this study were required to know the participant for at least 10 years. Other limitations include the over representation of women and more highly educated people in our sample and, as with most genetic studies, these results may not be generalizable to non-European ancestry populations. We note our sample size is relatively small compared with much of the wider genetic literature and replication in independent cohorts is needed. At present we are not aware of any other large cohort studies which use the MBI-C, however we would argue that our findings, along with the practical advantages of the MBI-C (it is freely available and quick to complete) provide a strong case the wider adoption of the scale which will allow important follow up work to take place.

## Conclusions

In summary, this study lends further support to the growing evidence base that later life emergent neuropsychiatric symptoms describe an at-risk state for incident cognitive decline and dementia, and can be the index manifestation of dementia for some, associated with dementia biomarkers and genetic risk. Our findings also support the case for using MBI as a sample enrichment tool for biomarker studies and clinical trials targeting at-risk individuals. The enrichment approach is inexpensive, simple, and scalable, and can decrease cost and improve enrolment efficiency of dementia clinical trials [45]. Deeper phenotyping of these groups including neuroimaging and longitudinal monitoring of clinical outcomes is now essential.

## Additional files

### Supplementary Material

File format: MS word.docx

Description: figures showing ancestry principal components

## Data Availability

The data that support the findings of this study are available from but restrictions apply to the availability of these data, which were used under license for the current study, and so are not publicly available. Data are however available from the authors upon reasonable request and with permission of the PROTECT study.

## Abbreviations

MBI: Mild Behavioral Impairment
NPS: neuropsychiatric symptoms
PRS: polygenic risk score
IQCODE: Informant Questionnaire on Cognitive Decline in the Elderly
IADL: instrumental activities of daily living

## Declarations

### Ethics approval and consent to participate

Ethical approval for the PROTECT study itself was obtained from the London Bridge Research Ethics Committee (reference 13/LO/1578). Written informed consent was obtained from all participants and proxy informants, this consent also covers secondary analysis of data by third party approval researchers so separate approval was not required for this analysis.

### Consent for publication

Not applicable.

### Authors’ contributions

BC: conceptualisation, analysis design, statistical analysis, interpretation of findings and manuscript drafting; ZI: conceptualisation, analysis design, interpretation of findings and manuscript drafting; CB: analysis design, interpretation of findings and manuscript drafting; HB: data acquisition, manuscript review; AC: data acquisition, manuscript review, funding; DA: data acquisition, manuscript review.

### Acknowledgements

This research was also supported by the NIHR Collaboration for Leadership in Applied Health Research and Care South West Peninsula. The views expressed are those of the author(s) and not necessarily those of the NHS, the NIHR or the Department of Health and Social Care. The authors thank Gemma Shireby at University of Exeter for providing the analysis script for genetic ancestry check. Genotyping was performed at deCODE Genetics.

